# Vapor H_2_O_2_ sterilization as a decontamination method for the reuse of N95 respirators in the COVID-19 emergency

**DOI:** 10.1101/2020.04.11.20062026

**Authors:** Ebru Oral, Keith K. Wannomae, Rachel Connolly, Joseph Gardecki, Hui Min Leung, Orhun Muratoglu, Anthony Griffiths, Anna N. Honko, Laura E. Avena, Lindsay G. A. McKay, Nick Flynn, Nadia Storm, Sierra N. Downs, Ralph Jones, Brandon Emmal

## Abstract

There are a variety of methods routinely used in the sterilization of medical devices using hydrogen peroxide (H_2_O_2_) including vaporization, plasma generation and ionization. Many of these systems are used for sterilization and are validated for bioburden reduction using bacterial spores.

Here, we explored the benefits of using vaporized H_2_O_2_ (VHP) treatment of N95 respirators for emergency decontamination and reuse to alleviate PPE shortages for healthcare workers in the COVID-19 emergency. The factors that are considered for the effective reuse of these respirators are the fit, the filter efficiency and the decontamination/disinfection level for SARS-CoV-2, which is the causative virus for COVID-19 and other organisms of concern in the hospital environment such as methicillin-resistant *Staphylococcus aureus* or *Clostridium difficile*. WE showed that the method did not affect fit or filter efficiency at least for one cycle and resulted in a >6 log reduction in bacterial spores and >3.8 log reduction in the infectious SARS-CoV2 load on N95 respirators.

## Background

Here, we explored the benefits of using vaporized H_2_O_2_ (VHP) treatment of N95 respirators for emergency decontamination and reuse to alleviate PPE shortages for healthcare workers in the COVID- 19 emergency. The factors that are considered for the effective reuse of these respirators are the fit, the filter efficiency and the decontamination/disinfection level for SARS-CoV-2, which is the causative virus for COVID-19 and other organisms of concern in the hospital environment such as methicillin-resistant *Staphylococcus aureus* or *Clostridium difficile*.

## Methods

N95 respirators (3M, 1860S; n=10) were processed in a VHP LTS-V (Steris, Mentor, OH). This is a standard sterilization cycle for medical devices where a sterility assurance level (SAL) 10^-6^ requirement is verified on device surfaces by using biological indicators (*Geobacillus stearothermophilus*) placed in the most challenging locations in the package/loads.

### 1. Respirator performance testing (Filter efficiency and fit)

Treated (n=5) and untreated respirators (n=3) were tested for filter efficiency (ICS Laboratories, Brunswick, OH) using an abbreviated test protocol ICS developed from NIOSH TEB-APR-STP-0059. After preconditioning for 24 hours at 38°C and 85 % relative humidity, filter media were challenged with NaCl aerosol at a flowrate of 85 L/min and a particle size of 0.075 CMD for both instantaneous and loading (200mg/m^3^ max) efficiencies. Qualitative fit testing of VHP treated respirators (n=4) was done with an aerosolized saccharin solution per OSHA § 1910.134 App A at MGH (Environmental Health and Safety) with volunteers (previously fit to this type of respirator).

### 2. Viral infectivity tests

All viral infectivity tests were performed at biosafety level 4 (BSL4) at the National Emerging Infectious Diseases Laboratories (NEIDL).

#### 2.1. Cells and virus

SARS-CoV-2 USA-WA1/2020 [1] was propagated in Vero E6 cells (BEI resources, NIAID, NIH: VERO C1008 (E6), African green monkey kidney, Working Bank #NR-596). Virus stock used for these studies had been passaged thrice in VERO cells and twice in VeroE6 cells. Cells were maintained in Dulbecco’s modified

Eagle medium (DMEM) with GlutaMAX™ and sodium pyruvate (Gibco™, ThermoFisher Scientific, Waltham, MA, catalog 10569), non-essential amino acids (Gibco™, catalog 11140) and 10% HI-FBS (Gibco™, catalog 10082).

#### 2.2. Decontamination of SARS CoV-2 contaminated N95 masks using VHP

N95 respirators were trimmed to ∼ 2 cm^2^ segments along the outer edge to ensure all layers remained intact and were transferred to the BSL4 laboratory. 50µL of SARS-CoV-2 was applied to each respirator segment, covering approximately half of the outer facing surface, and allowed to air dry in a biosafety cabinet (BSC) for approximately 2 hours. As a control, virus was applied to plastic tissue culture dishes (Corning™ CellBIND™, catalog 3335); one was exposed to VHP treatment and one was retained at ambient temperature. To examine the effects of VHP treatment on subsequent SARS-CoV-2 contamination, a set of respirators were treated with VHP, followed by virus application and drying. These groups are represented below in Table 1.

**Table 1.**
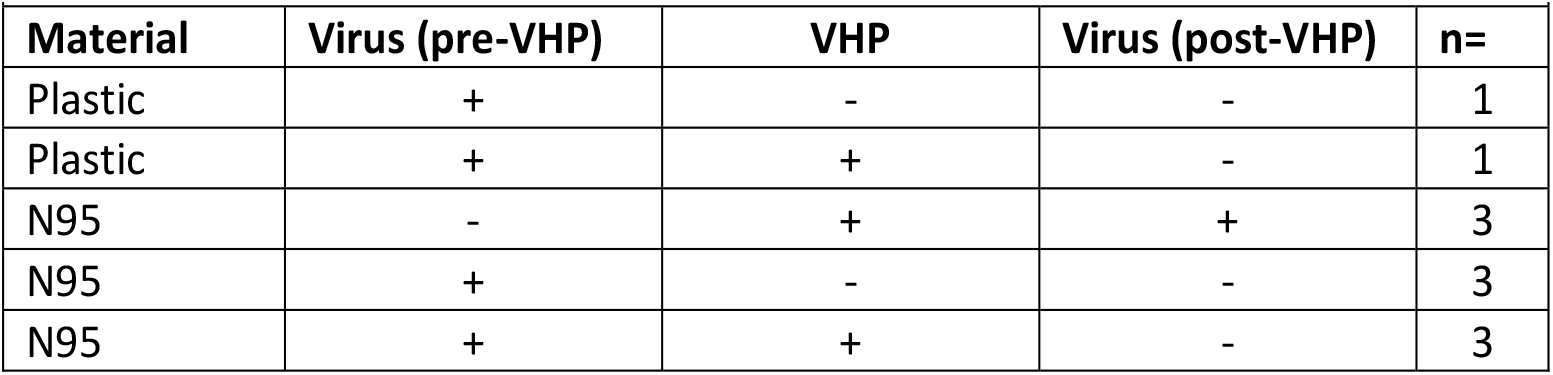
Sample groups.

| Table 1. Sample groups. |  |  |  |  |
| --- | --- | --- | --- | --- |
| Material | Virus (pre-VHP) | VHP | Virus (post-VHP) | n= |
| Plastic | + | - | - | 1 |
| Plastic | + | + | - | 1 |
| N95 | - | + | + | 3 |
| N95 | + | - | - | 3 |
| N95 | + | + | - | 3 |

The VHP treatment was performed using a Steris ARD1000^®^ in a 691 ft3 room; samples were exposed to 410± 83 ppm of vaporized hydrogen peroxide for approximately 3 hours followed by off-gassing until hydrogen peroxide was undetectable (0 ppm, approximately 4.5 hours total from initiation of VHP cycle). Following VHP exposure, respirator pieces were added to 2mL of DMEM + 2% FBS + antibiotics/antimycotics (Gibco™) and vortexed at 2,000 rpm for 2 minutes, followed by centrifugation at 1,000 RCF for 5 minutes. Virus from plastic dishes was resuspended in 2mL of complete DMEM. Infectivity was quantified using a plaque assay in duplicate using 200 µL of the undiluted eluate to a 1:10,000 dilution. Additionally, samples were plated (500µL) in duplicate on cells and following a 1-hour incubation, 1.5mL complete DMEM was added. These plates were observed for cytopathic effect for five days.

#### 2.3. Virus titration by plaque assay

Samples were diluted in DMEM and 10% HI-FBS (Gibco™) with Antibiotic-Antimycotic (Gibco™) and 200µL added to confluent monolayers of NR-596 Vero E6 cells in duplicate. Samples were adsorbed to the cell monolayers for 1 h at 37 °C and 5% CO_2_ with gentle rocking approximately every 15 minutes to prevent monolayer drying. The monolayers were then overlaid with a 1:1 mix of 2.5% Avicel^®^ RC-591 microcrystalline cellulose and carboxymethylcellulose sodium (DuPont Nutrition & Biosciences, Wilmington, DE) mixed with 2X Modified Eagle Medium (Temin’s modification, Gibco™) supplemented with 2X Antibiotic-Antimycotic (Gibco™), 2X GlutaMAX (Gibco™) and 10% HI-FBS (Gibco™). Cells were incubated at 37 °C and 5% CO_2_ for 2 days, fixed with 10% neutral buffered formalin before removing from containment, then stained with 0.2% aqueous Gentian Violet (RICCA Chemicals, Arlington, TX) in 10% neutral buffered formalin for 30 min, rinsed, and plaques were enumerated.

## Results

### Respirator performance testing

The VHP cycle did not compromise the filter efficiency of 3M, 1860S N95 respirators (Table 2). All four respirators passed fit testing on human subjects.

**Table 2.** The effects of vaporized hydrogen peroxide (VHP) decontamination treatment on the fit and filter efficiency of the N95 respirators.

|  |  |  |  |  |
| --- | --- | --- | --- | --- |
| Samples | Number Tested | Penetration of particles (%) | Breathing resistance (mmH<sub>2</sub>O) | Fit test |
| Untreated | 3 | 0.8 ± 0.2<br>(0.64 – 0.95) | 11.5 ± 0.7<br>(10.93 – 12.26) |  |
| VHP Treated (Steris LTS-V) | 5 | 1.2 ± 0.2<br>(0.99 – 1.50) | 10.0 ± 0.6<br>(9.27 – 10.53) | Pass (4/4) |

### SARS CoV-2 inactivation on face masks

VHP treatment of respirators intentionally contaminated with SARS-CoV-2 reduced the infectious virus load to below the limit of detection (1.3 log_10_, which is calculated based on the volumes of eluate used in the experiments and the premise that 1 PFU could be detected by the assays; Table 3 and Figure 1). There were no plaque forming units for samples treated with VHP. This corresponded to >3.8 log_10_ reduction in infectious virus load compared to the virus stock and a >2.6 log_10_ reduction compared to the infectious virus load on the respirators.

**Table 3.** Infective SARS-CoV2 concentration of study samples.

| Table 3. Infective SARS-CoV2 concentration of study samples. |  |
| --- | --- |
| Treatment | SARS-CoV-2 (PFU per sample) |
| Virus Stock | 1.24 x 10<sup>5</sup> |
| Virus on Plastic (no VHP) | 5.00 x 10<sup>3</sup> |
| Virus on Plastic + VHP | below detection limit |
| Virus + N95 (no VHP) | 7.85 x 10<sup>3</sup> |
| Virus + N95 + VHP | below detection limit |
| N95 + VHP (Virus after) | 4.70 x 10<sup>3</sup> |

**Figure 1:**
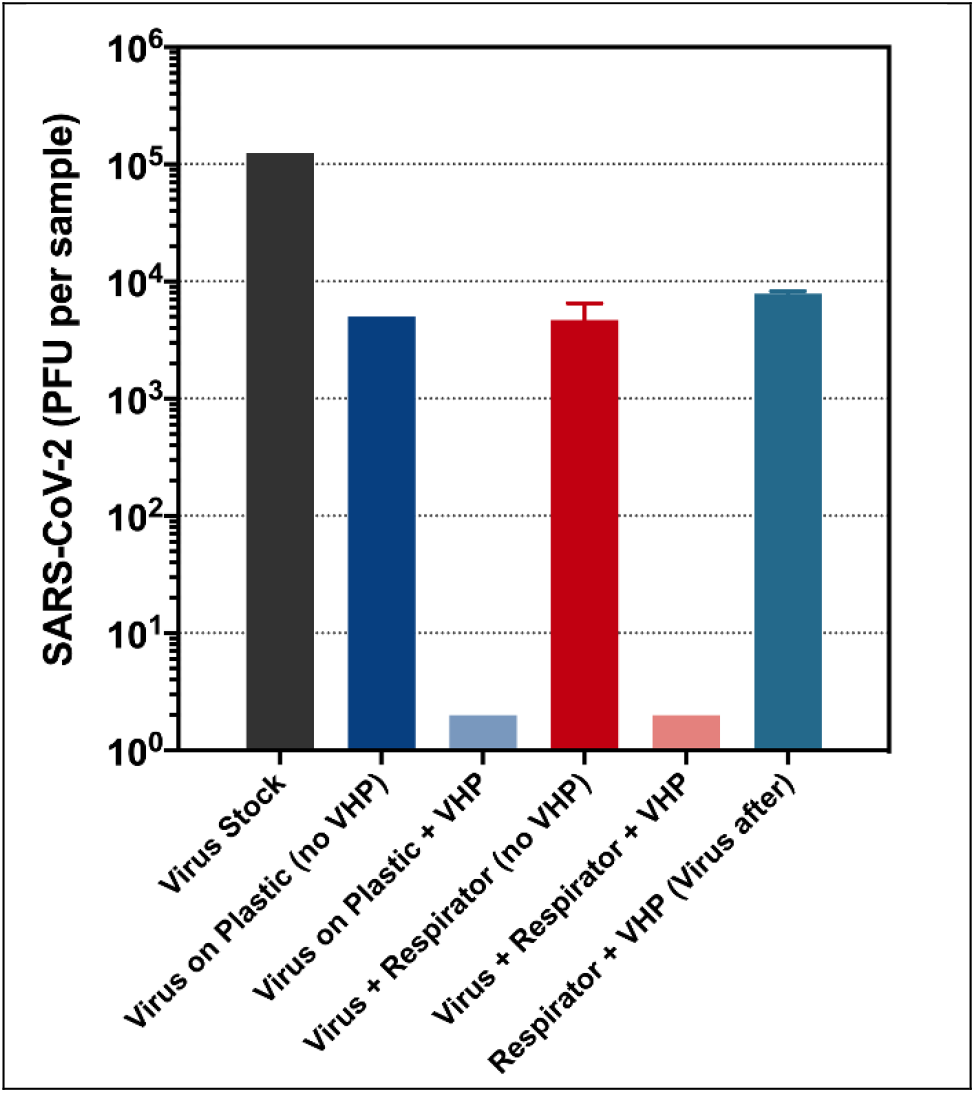
The effect of various treatments on the infectivity of SARS CoV-2

## Discussion

In this study, one standard cycle of VHP sterilization (Steris LTS-V) for one type of N95 respirator was found to be feasible in terms of preserving fit and filter efficiency. Returning the respirators to the index users after decontamination for reuse may ensure fit for a longer period. Testing is ongoing to determine how the fit and filter efficiency are affected by multiple decontamination and reuse cycles. A VHP system provided by the Battelle Memorial Institute (https://www.fda.gov/media/136529/download) and several low temperature sterilization systems by Steris (https://www.fda.gov/media/136843/download) have obtained an Emergency Use Authorization for this application. These systems are approved for use for 20 and 10 decontamination cycles, respectively.

One concern with VHP as with the use of other gas sterilant is the residuals that may build up on the respirators and may be inhaled by the end-user. A detailed analysis of pentane extracted respirators (S1) treated with VHP by Salter et al. followed by the prediction of inhaled values compared to safety levels concluded that there was no significant threat to the healthcare worker in a realistic scenario after at least one cycle [2].

The concentration and time of hydrogen peroxide exposure in both units tested here are designed for sterility assurance level of 10^-6^ (6-log_10_ reduction) with *Geobacillus stearothermophilus* spores. Typically, process validation using these spores for the application/load is done initially and at regular intervals for a system and verified with chemical indicators in each run. Both the LTS-V system and the ARD1000 setup used for viral infectivity studies were validated by bacterial spores. Thus, H_2_O_2_ exposure under these conditions provided 6-log bioburden reduction of bacterial spores, which are the most resistant microorganisms. A ‘high level of disinfection’, also against common clinical pathogens such as Staphyloccocal species and *C. difficile*, is expected as indicated in the FDA guidance entitled “Enforcement Policy for Face Masks and Respirators During the Coronavirus Disease (COVID-19) Public Health Emergency (Revised)” (April 2020). In this document, the FDA also recommends the demonstration of viricidal activity wherever possible (≥ 3-log).

A total of 1.24×10^5^ PFU (5.09 log_10_) in 50 µL was inoculated on to respirators that were treated with VHP and no infectious virus could be detected in the eluate following treatment. This includes plaque assays that used 200 µL of eluate (Figure 2) and plating 500 µL of eluate followed by monitoring the cells for cytopathic effect for five days to improve sensitivity. These results suggest that VHP treatment was effective in inactivating SARS-CoV2.

**Figure 2:**
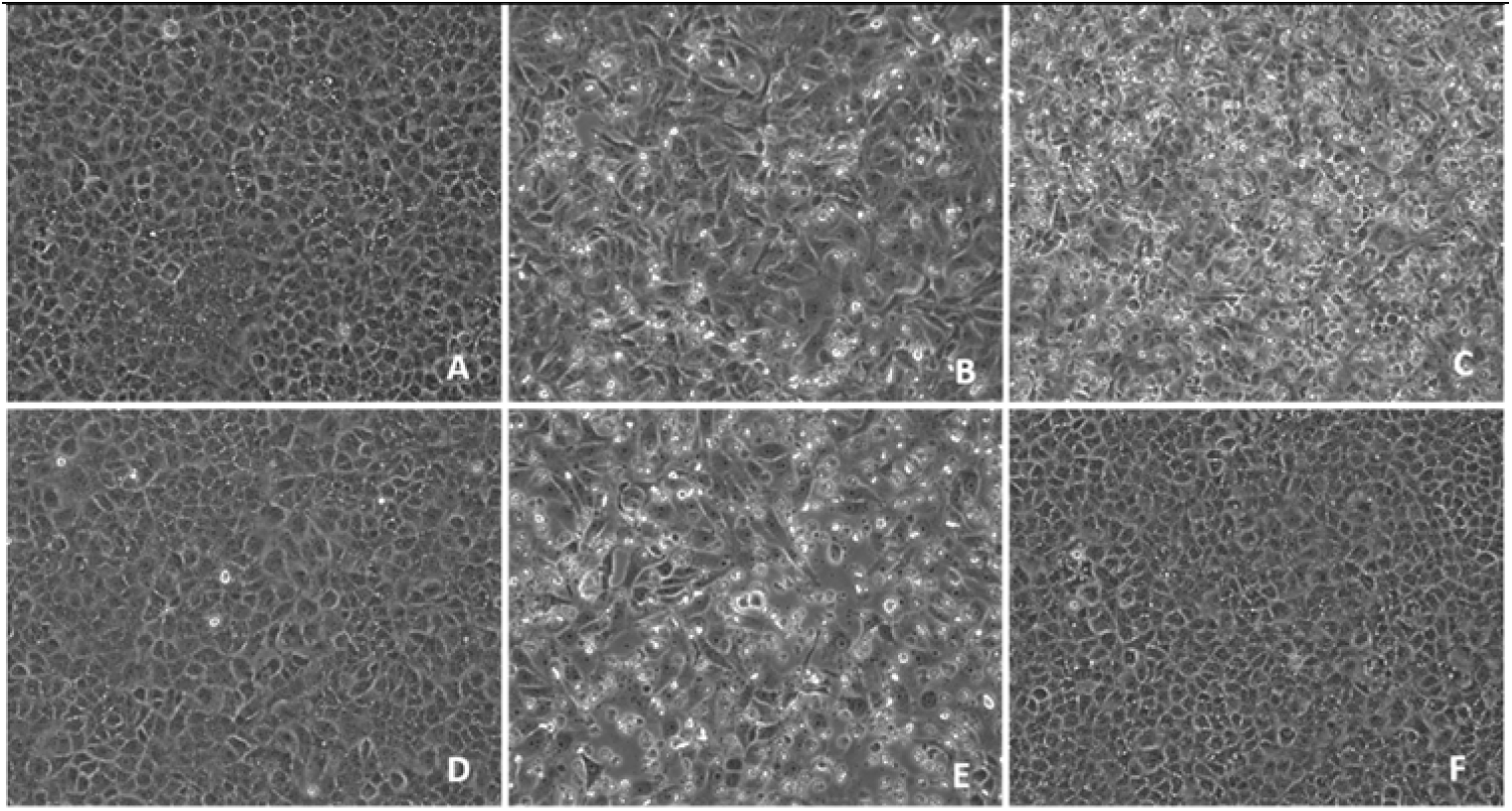
Cytopathic effect (CPE) in NR-596 Vero E6 cells following vaporized hydrogen peroxide (VHP) decontamination treatment (images are 20X magnification). No CPE was observed in mock-inoculated cell monolayers, or in cell monolayers inoculated with virus treated with VHP or eluted from respirators treated with VHP. A) Mock-inoculated cell monolayer B) CPE on cell monolayer inoculated with virus recovered from a VHP-treated respirators (virus added to respirator after treatment). C) Virus recovered from respirator not treated with VHP. D) Virus recovered from VHP-treated respirator (virus added to respirator before treatment). E) Virus on plastic not treated with VHP. F) Virus on plastic treated with VHP.

Of the 1.24×10^5^ PFU that were loaded onto masks, 7.85 × 10^3^ PFU (3.89log_10_) could be eluted. The approximately 16-fold reduction (1.2 log_10_) in infectivity is due to a combination of viral inactivation due to contact with the respirator and the inability to elute all infective virus from the respirator. At the same time, contact with the tissue culture flask decreased infectivity similarly, suggesting that the recovery from the respirator was fairly efficient. The respirators laden with virus after VHP treatment of the respirator also showed a similar amount of infective virus load (4.70×10^3^ PFU were eluted from a respirator previous treated with VHP). This further suggested that there were no residuals on the respirators after VHP treatment that interfered with viral infectivity.

A limitation of the experiment is the static application of virus to the respirators unlike the clinical use cases where there is active air flow through the respirator via inhalation by the healthcare worker. This may enable the penetration of the aerosolized droplets containing the virus deeper into the respirator. Further study of the infective virus load in clinically used respirators can determine if this is a factor to be considered for decontamination methodology.

While we present results for the technical and bioburden evaluation of this decontamination method for N95 respirators, there are many other considerations such as financial, and logistical ones for any medical institution to decide on a decontamination method, if any, to utilize for this emergency use situation. The results presented here are not meant to constitute a stand-alone recommendation.

## Data Availability

This is non-clinical, esperimental study. The data are presented in the preprint. Any other details can be accessed by contacting the corresponding author.

## Acknowledgements

We would like to acknowledge CONFORMIS (Billerica, MA) for allowing us the use of their Steris LTS-V sterilizer, and Dr. Galit Alter for valuable discussions on viral infectivity. Avicel^®^ RC-591 was provided for these efforts by DuPont Nutrition & Biosciences, Wilmington, Delaware.

